# Naturally acquired Rift Valley fever virus neutralizing antibodies predominantly target the Gn glycoprotein

**DOI:** 10.1101/2020.08.07.20170241

**Authors:** Daniel Wright, Elizabeth R. Allen, Madeleine H.A. Clark, John N. Gitonga, Henry K. Karanja, Ruben Hulswit, Iona Taylor, Sumi Biswas, Jennifer Marshall, Damaris Mwololo, John Muriuki, Bernard Bett, Thomas A. Bowden, George M. Warimwe

## Abstract

Rift Valley fever (RVF) is a viral haemorrhagic disease first discovered in Kenya in 1930. Numerous animal studies have demonstrated that protective immunity is acquired following RVF virus (RVFV) infection, and that this correlates with acquisition of virus neutralizing antibodies (nAb) that target the viral envelope glycoproteins. However, naturally acquired immunity to RVF in humans is poorly described. Here, we characterized the immune response to the viral envelope glycoproteins, Gn and Gc, in RVFV-exposed Kenyan adults. Long-lived IgG (dominated by IgG1 subclass) and T cell responses were detected against both Gn and Gc. However, antigen-specific antibody depletion experiments showed that Gn-specific antibodies dominate the RVFV nAb response. IgG avidity against Gn, but not Gc, correlated with nAb titers. These data are consistent with the greater level of immune accessibility of Gn on the viral envelope surface and confirm the importance of Gn as an integral component for RVF vaccine development.

## INTRODUCTION

Rift Valley fever (RVF) is one of several epidemic diseases prioritised by the World Health Organization and other global organisations for urgent research and development of control interventions (Mehand et al., 2018, Gouglas et al., 2018). The disease was first described in Kenya in 1930 but RVF epidemics are now commonly reported in Africa and the Arabian Peninsula (CDC, 2020). RVF is caused by the mosquito-borne Rift Valley fever virus (RVFV), a single-stranded RNA *Phlebovirus* in the *Phenuviridae* family (ICTV, 2020). The virus primarily infects ruminants resulting in high rates of neonatal mortality and abortion, that translate to major economic losses in the livestock industry of affected countries. Zoonotic transmisson to humans most commonly occurs during livestock epizootics through infectious mosquito bites or direct contact with tissue or fluid from infected animals (Daubney et al., 1931a, Nicholas et al., 2014). A wide spectrum of clinical manifestations characterize RVF in humans; including self-limiting mild illness associated with fever, myalgia and other non-specific symptoms, through to haemorrhagic and neurological manifestations with high case fatality and debilitating sequela, and miscarriage in pregnant women (reviewed in (Wright et al., 2019)). No therapeutics or vaccines are currently approved for human use, though several candidate vaccines are in development (Gouglas et al., 2018).

RVFV contains a tripartite genome consisting of a small (S), medium (M) and large (L) segment (Wright et al., 2019). The S segment encodes the nucleoprotein (N) and a non-structural protein (NSs). The L segment encodes the RNA-dependant RNA polymerase. The M segment encodes the structural glycoproteins, Gn and Gc. These two glycoproteins form heterodimers arranged into higher order pentamers and hexamers that encapsulate the mature virion and are responsible for host-cell attachment and membrane fusion (Halldorsson et al., 2018a, Freiberg et al., 2008, Sherman et al., 2009, Huiskonen et al., 2009). They are the prime targets of neutralizing antibodies. Through the use of alternative translation sites and post-translational cleavage, the M segment also encodes for a non-structural protein and a 78kDa protein, which may play a structural role in virus-derived from mosquito cells (Weingartl et al., 2014).

Historical seminal studies in livestock and non-human primates demonstrated that immunity to RVF is acquired following recovery from RVFV infection, and that this can be passively transferred to susceptible animals through administration of convalescent sera (Peters et al., 1988, Daubney et al., 1931b). These studies identified viral neutralizing antibodies (nAb), which target the RVFV Gn and Gc envelope glycoproteins, as the main correlates of protection against RVF (Easterday, 1965). Occupational exposure to RVFV infection during the decades following the discovery of RVFV provided the first evidence of naturally acquired RVFV nAbs and these tended to be long-lived, being readily detectable in two individuals 12 and 25 years post-infection, respectively, despite no further exposure (Smithburn et al., 1949, Brown et al., 1957, Findlay, 1936). However, beyond these studies, naturally acquired immunity to RVFV in humans is poorly described. Here, we sought to address this paucity of information by characterizing the humoral and cellular response to RVFV among naturally exposed adults in Kenya, including an investigation of the longevity and kinetics of RVFV nAbs in this population.

## RESULTS

Stored samples from two adult populations in coastal Kenya were used for this study (see Methods). One population of adults comprised a random selection of fifty RVFV-exposed individuals included in a previous cross-sectional survey investigating the risk factors for RVFV exposure in Tana River county (Bett et al., 2018). The second population was composed of adults under longitudinal surveillance for malaria within the KHDSS in Kilifi county and had previously not been screened for RVFV exposure (see Methods).

### IgG responses against Gn and Gc correlate with RVFV nAb levels

We first measured IgG antibody responses against Gn and Gc in fifty RVFV-exposed adults from Tana River county and assessed their relationship with the RVFV nAb response. The geometic mean titer (GMT) of RVFV nAb as measured by virus neutralizing assay (VNT_50_) in these fifty adults was approximately 2100 (95% CI 1543, 2792), and ranged between ~160 to >14000 (**Fig. 1**). Total IgG titers against Gn and Gc (ELISA GMT of 67.3 for Gn and 66.9 for Gc) were highly correlated (r=0.77, p<0.001). A strong correlation was observed between the RVFV nAb titer and IgG response against Gn (r=0.76, p<0.0001) and Gc (r=0.68, p<0.0001) (**Fig 1a**). IgG1 subclass predominated the antibody response against both Gn and Gc (**Fig. 1c**).

**Figure 1.**
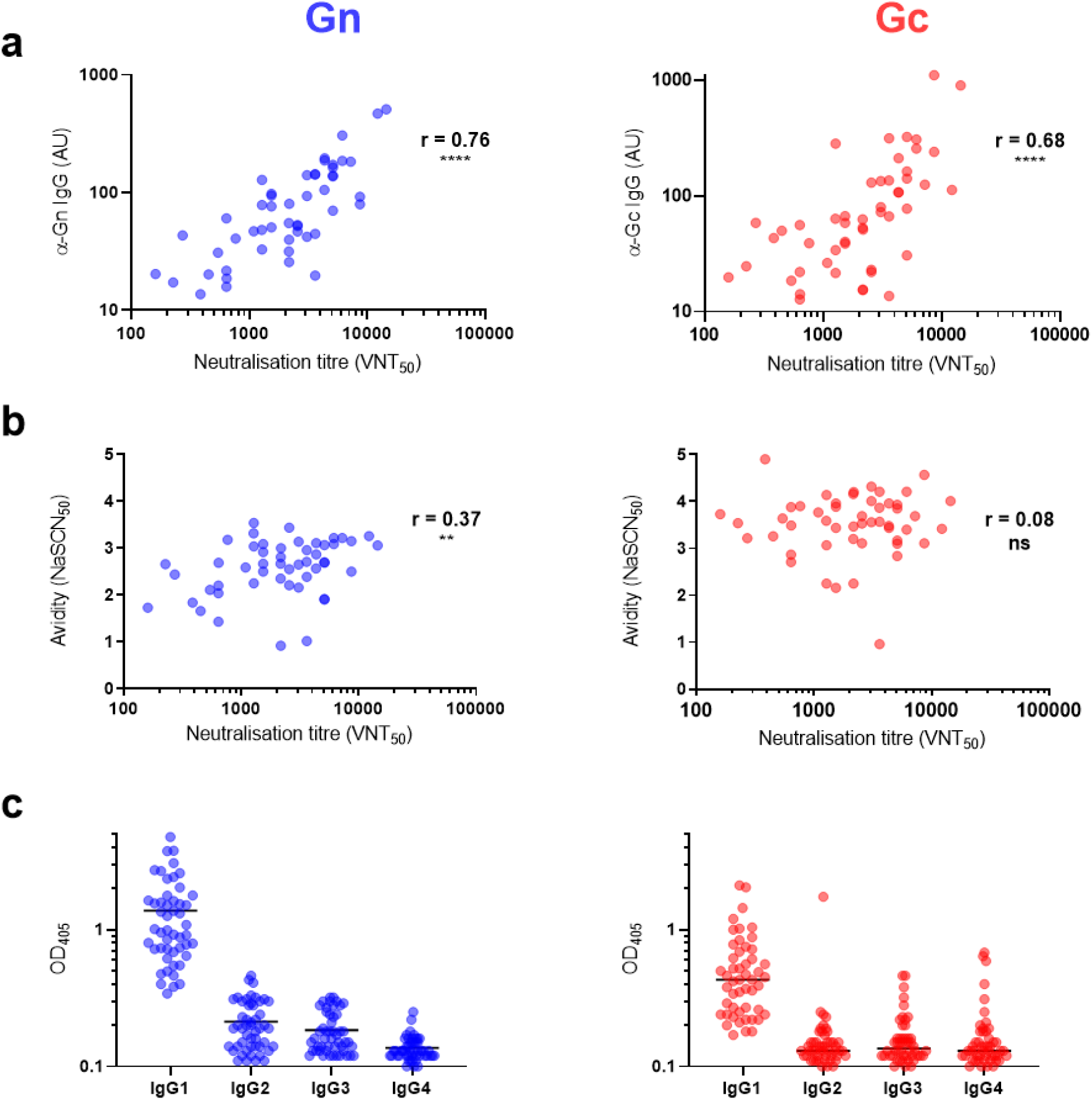
*Total IgG titer towards recombinant Gn and Gc correlate strongly with neutralizing antibody titer. (**a**) Correlations between neutralization titer, as measured by VNT*_50_ *assay, and total IgG titer towards Gn and Gc, as measured by ELISA (Gn: r=0.76, 95% CI: 0.60-0.86. Gc; r=0.68, 95%CI: 0.49- 0.81).**** = P<0.0001. (**b**) Correlations between neutralization titer and avidity of total IgG towards Gn and Gc (Gn: r=0.37, 95% CI: 0.09-0.59, **; Gc: r= 0.08, 95%CI: -0.21-0.36, p=0.57).** = P=0.009. (**c**) IgG subclass response towards Gn and Gc. Lines represent geometric means*.

We also measured the avidity of the antibody response to each glycoprotein, expressing this as the concentration of NaSCN, a chaotropic agent, required to reduce ELISA signal by 50% (NaSCN IC50). Anti-Gc antibody avidity was significanty higher than that for anti-Gn antibodies (**Fig. 1b**); however, a correlation between antibody avidity and nAb response was only observed for Gn (r=0.37, p=0.009) and not Gc (r=0.08, p=0.57).

### nAbs from naturally exposed adults preferentially target RVFV Gn over Gc

We next investigated the relative contribution of Gn and Gc to the RVFV nAb response. To do this, we pooled sera from 7 RVFV-exposed adults from Tana River county that had sufficient material available. We then depleted the pooled serum of antibodies specific for one or both of the recombinant RVFV glycoproteins (see Methods). ELISA was used to confirm the depletion, showing a reduction in IgG endpoint titer of 98% and 93% for Gn and Gc, respectively, compared to non-depleted serum (**Fig. 2a**). The impact of this antigen-specific antibody depletion on the ability of the pooled serum to neutralize RVFV *in vitro* was then measured by VNT_50_. Anti-Gn antibody depletion resulted in a 79% reduction in nAb titer compared to the non-depleted serum (**Fig. 2b**). In contrast, depletion of anti-Gc antibodies from sera showed a modest 29% reduction compared to the non-depleted serum (**Fig. 2b**). Depletion of both Gn and Gc antibodies showed a 79% reduction in nAb titer, identical to the pool depleted of anti-Gn antibodies alone (**Fig. 2b**).

**Figure 2.**
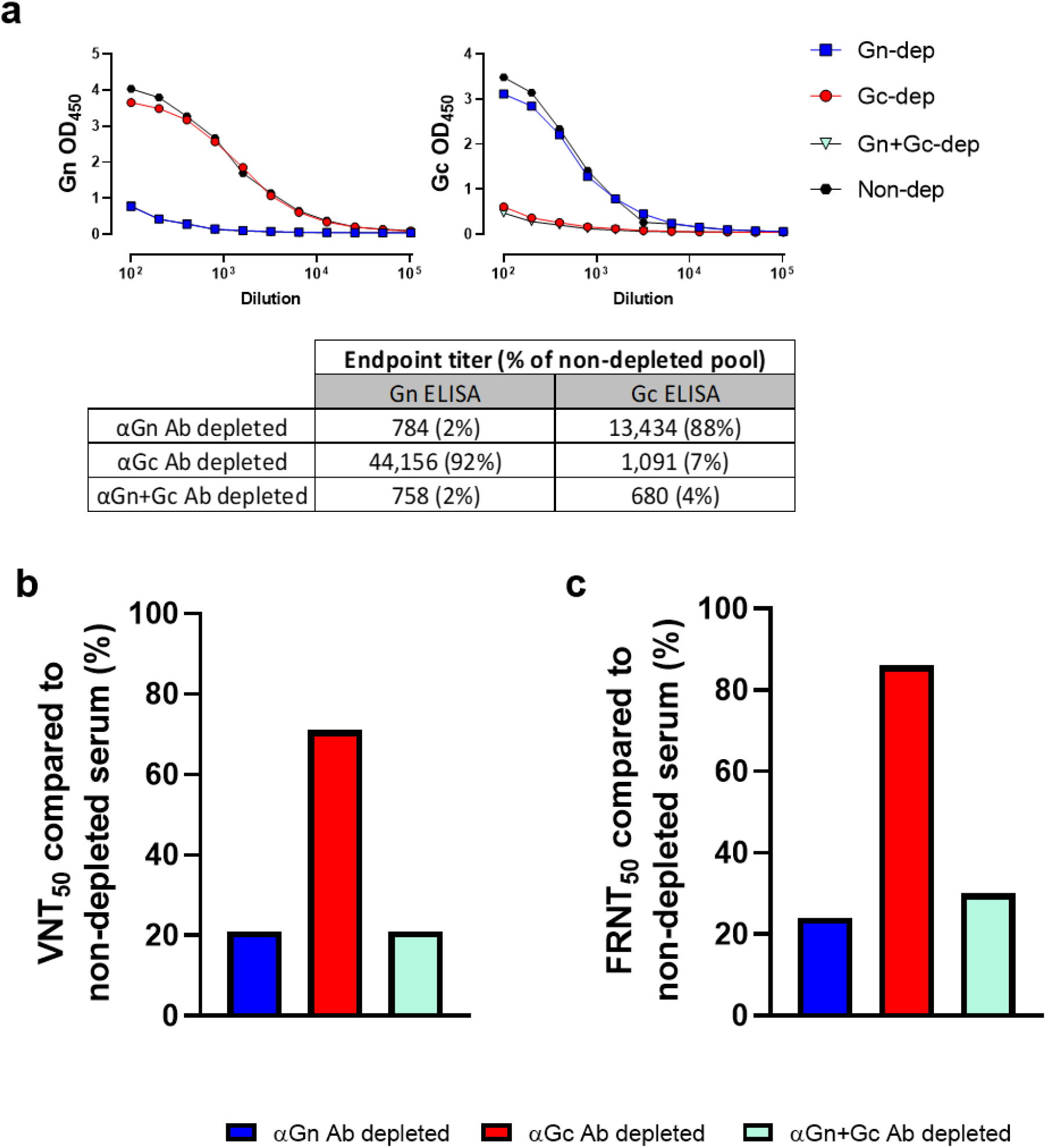
*Neutralizing antibody titers predominantly target RVFV Gn over Gc. Individual sera were pooled from individuals in Tana River county (n=7) and depleted of anti-Gn antibodies, anti-Gc antibodies, both or neither. (a) Confirmation of depletion by Gn ELISA (left) and Gc ELISA (right) with Gn and Gc Endpoint titers of each pool calculated in the table. (b) Neutralization titer (VNT*_50_*) of each depleted pool as a percentage of non-depleted serum (VNT*_50_ *of Gn antibody depletion, 1600; Gc antibody depletion, 5344; Gn+Gc antibody depletion, 1600; non-depleted, 7557). (c) Neutralization titer (FRNT50) after repeating experiment using an entirely different serum pooled from individuals from Kilifi county (n=6)*.

Of 200 adults under longitudinal surveillance for malaria studies in Kilifi county, 6 (3%) in 2018 had evidence of RVFV exposure on the basis of carriage of RVFV nAbs (see Methods). To confirm the predominance of Gn as a target of the RVFV nAb response, we pooled sera from these 6 individuals and repeated the depletion experiment described above and used a focus reduction neutralization assay (FRNT50) to measure RVFV nAbs (see Methods). Results were consistent, showing a relative reduction in FRNT50 of approximately 76%, 14% and 70% in sera depleted of antibodies to Gn, Gc or both, respectively (**Fig. 2c**). These data support RVFV Gn as the major target of the nAb response arising during natural RVFV infection.

### IgG and RVFV nAb titers remain high over many years

To study longevity of the observed immune respones we used three of the six RVFV-exposed adults from Kilifi county that had corresponding stored serum samples spanning multiple years (see Methods). We also included stored sera from an additional two RVFV-exposed adults identified from the same surveillance study in previous years. Overall, the period covered by each individual’s samples ranged from 3-11 years, but dates of RVFV exposure were unknown (**Fig. 3**). Both IgG (anti-Gn and anti-Gc) and RVFV nAb titers were readily detected over the period of follow up (**Fig. 3**). In keeping with the two seminal studies on the durability of RVFV nAb titers, the individual from whom we had the highest number of samples, JA0073 (n=8), exhibited high nAb titers over an 11-year period (**Fig. 3a**). RVFV nAb titers were generally stable with only slight reductions from individual peak nAb titers observed. One individual, JA0193, had a >12 fold increase in nAb titer between their baseline sample and the sample collected 5 years later, indicating RVFV re-exposure in the intervening period. Total binding IgG titers towards both glycoproteins were also long-lasting, remaining high despite a gradual decline over the years, while avidity remained stable (**Fig. 3b, c**).

**Figure 3.**
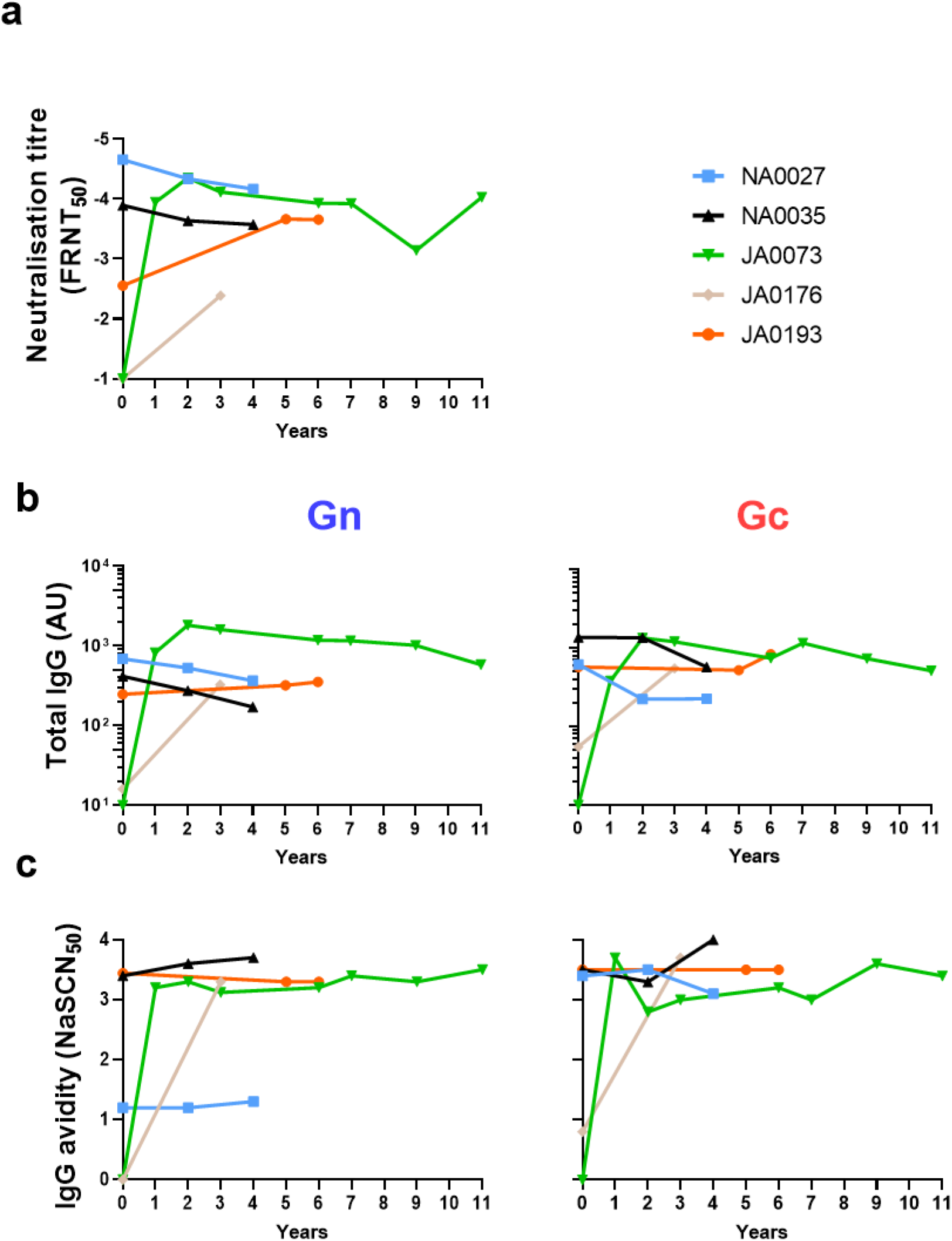
*Natural exposure to RVFV elicits a high-titer, long-lived humoral response. Five indivudals from Kilifi county (see text) with serum samples spanning 3-11 years were assayed for (**a**) Neutralization titer, (**b**) IgG titer of anti-Gn (left) and anti-Gc (right), (**c**) IgG avidity of anti-Gn IgG (left) and anti-Gc IgG (right). Year 0 is their earliest available sample, not necessarily the year of exposure, with subsequent samples plotted relative to that*.

### RVFV infection elicits a moderately low but significant cellular response to Gn and Gc

Cryopreserved PBMCs were also available from 5 of the RVFV-exposed adults sampled in 2018 from Kilifi county, allowing an assessment of RVFV-specific T cell responses. We measured Gn and Gc-specific IFNγ responses by ELISpot in five individuals and compared these to PBMCs from five randomly selected individuals from the same population who had no evidence of RVFV exposure (on the basis of lacking RVFV nAb or Gn/Gc IgG responses). Overall, the IFNγ response was moderately low in the five RVFV-exposed individuals (mean response 67.4 SFC/10^6^ PBMC 95% CI 6.8-128 SFC/10^6^) but this was significantly higher than non-exposed controls (mean response 8.4 SFC/10^6^ PBMC 95% CI 0-21.7 SFC/10^6^)(**Fig. 4**). Responses among RVFV-exposed individuals were comparable across individual peptide pools (**Fig. 4**). Staphylococcal enterotoxin B (SEB) was used as a positive control for PBMC stimulation and all unexposed and RVFV-exposed individuals had a response >1900 SFC/10^6^ PBMC as expected. Together, these data indicate that natural RVFV exposure elicits a durable T cell response.

**Figure 4.**
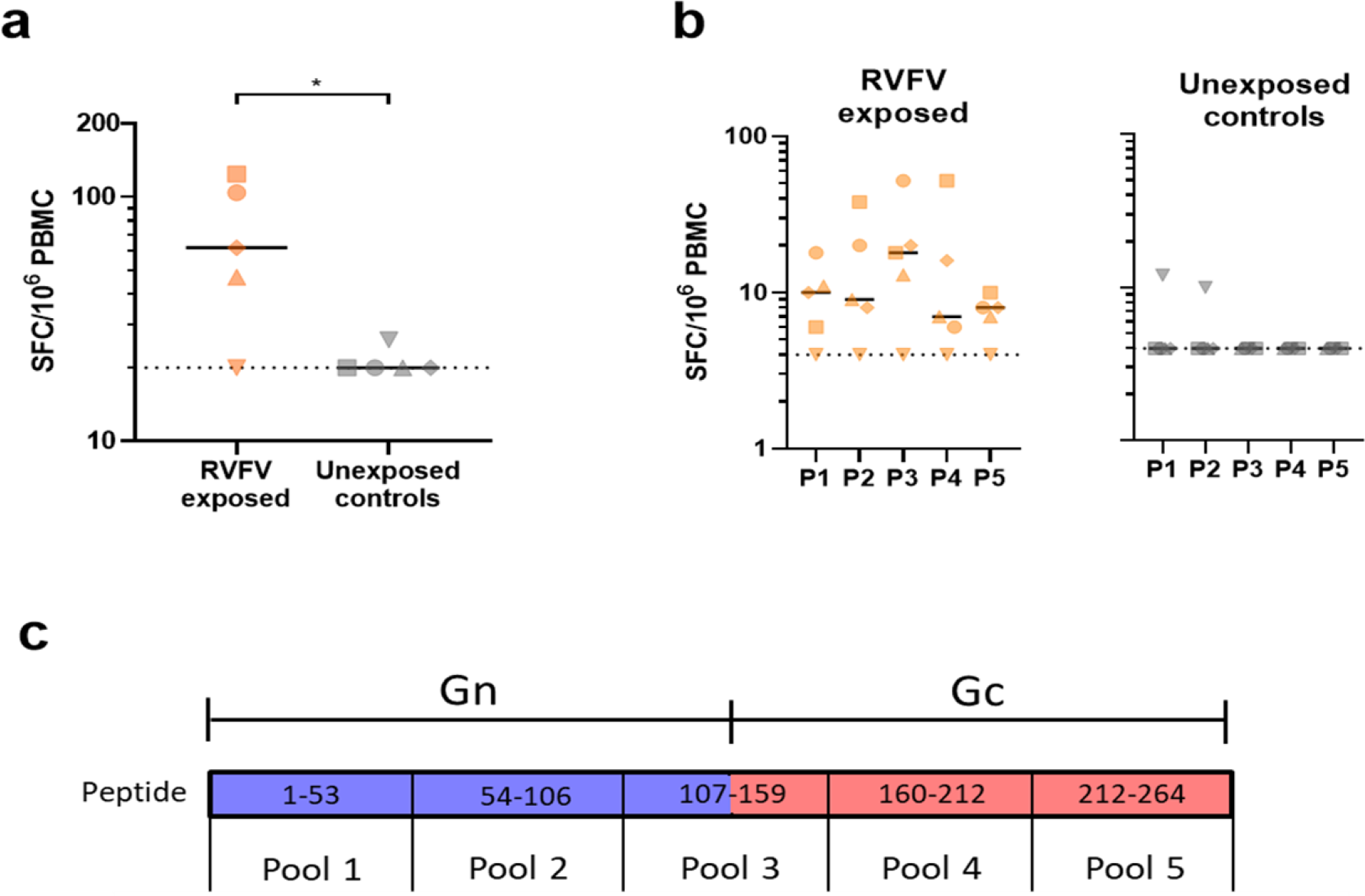
*A low but significant Ex vivo IFN*γ *ELISpot response in naturally exposed individuals. PBMC from five RVFV-exposed individuals along with 5 case-matched non-exposed controls were stimulated with an overlapping peptide library spanning the GnGc polyprotein. (a) Summed responses from the five peptide pools showing significantly higher response in the naturally exposed group compared to naive controls (*P<0.05, unpaired t test). (b) Responses to individual peptide pools, dotted line represent the lower limit of detection of 4 SFC/10^6^ PBMC for individual pools and 20 SFC/10^6^ PBMC for summed responses. SEB stimulated cells for each volunteer gave >1900 SFC/10^6^ PBMC (c) Schematic of the overlapping peptide library used for stimulation*.

## DISCUSSION

The protective role antibodies play against RVFV infection is well documented (Peters et al., 1988, LaBeaud, 2010, Niklasson et al., 1984). The structural glycoproteins displayed on the surface of RVFV, Gn and Gc, are key targets of this protective immune response and monoclonal antibodies have been identified targeting both (Besselaar and Blackburn, 1991, Besselaar-Blackburn, 1992, Wang et al., 2019). However, relatively little is known about the nature and longevity of the immune response after natural RVFV exposure. Here, using samples from adults in coastal Kenya with evidence of previous exposure to RVFV we show that: i) RVFV Gn and Gc glycoproteins are both targets of the long-lived T cell and IgG response to RVFV, and that the antibody response is predominated by the IgG1 sub-class, ii) RVFV nAb titers strongly correlate with Gn and Gc-specific IgG responses, and that iii) Gn is the main target of the RVFV nAb response. We also found that IgG avidity against Gn (but not Gc) correlated with the RVFV nAb response, further highlighting qualitative differences in the antibody response to both glycoproteins.

Studies in mice and rabbits have identified monoclonal antibodies targeting either Gn or Gc that are able to neutralize virus and confer protection against RVFV challenge (Besselaar and Blackburn, 1991, Besselaar-Blackburn, 1992, Keegan and Collett, 1986, Allen et al., 2018). Initial evidence of the possible greater role that antibodies targeting Gn play in protection, with respect to the Gc, came from reports involving passive transfer of anti-Gn or anti-Gc antibodies in mice, with only anti-Gn providing protection (Battles and Dalrymple, 1988). Another study in mice, this time involving vaccination with Gn-or Gc-coding DNA, showed that those vaccinated with Gn, but not Gc, developed nAbs (Lagerqvist et al., 2009). A more recent study isolating mAbs from a convalescent human patient returning to China found that mAbs specific to Gn exhibited much higher neutralization titers than those that bind Gc (Wang et al., 2019). This provides some evidence, perhaps, that the larger negative impact on neutralization titer seen in our study after depleting antibodies targeting Gn, compared to Gc, reflects a greater neutralizing potential of anti-Gn antibodies rather than merely increased abundance over anti-Gc antibodies. Further evidence supporting this hypothesis can be observed from our measurement of IgG avidity. Higher avidity antibodies can bind antigen more efficiently, and only the higher avidity of anti-Gn antibodies was associated with increased nAb titer. While avidity was very stable over many years, it is typically lower in acute or recent infection. However, due to the sparse sampling (no more than once a year), initial changes in the weeks and months post-exposure are not visible.

It is important to consider the differences between monomeric and soluble RVFV Gn and Gc used in these assays and their higher order membrane-bound pentameric and hexameric assemblies on the virion surface (Lozach et al., 2011, Leger et al., 2016). While our data shows that dual depletion of both Gn and Gc antibodies reduces nAb titers to identical levels seen in sera depleted of Gn antibodies alone, even dual-depleted sera can neutralize live virus at the highest concentration (**Fig. 2**). While this is likely due, at least in part, to the incomplete nature of the depletion (corresponding ELISA signal post-depletion was not entirely eliminated at high serum concentrations), it is also likely that a subset of antibodies recognize epitopes constituting higher-order quaternary assemblies formed on the native virion surface. These antibodies may be capable of neutralizing whole virus but may not bind monomeric Gn or Gc and therefore would not have been depleted. Nevertheless, we were still able to assess the relative contribution antibodies targeting each individual glycoprotein have on neutralization. While both glycoproteins are clearly immunogenic and targeted by the nAb response, our data suggest that after naturally acquired human infection antibodies targeting Gn, rather than Gc, play a larger role in RVFV neutralization. This is perhaps not surprising considering that the N-terminal component of Gn occupies the most membrane-distal region of the virus, which plays a role in shielding the cognate Gc (Halldorsson et al., 2018a, Allen et al., 2018), and therefore may be more easily accessible to the host immune system (**Fig. 5**).

**Figure 5.**
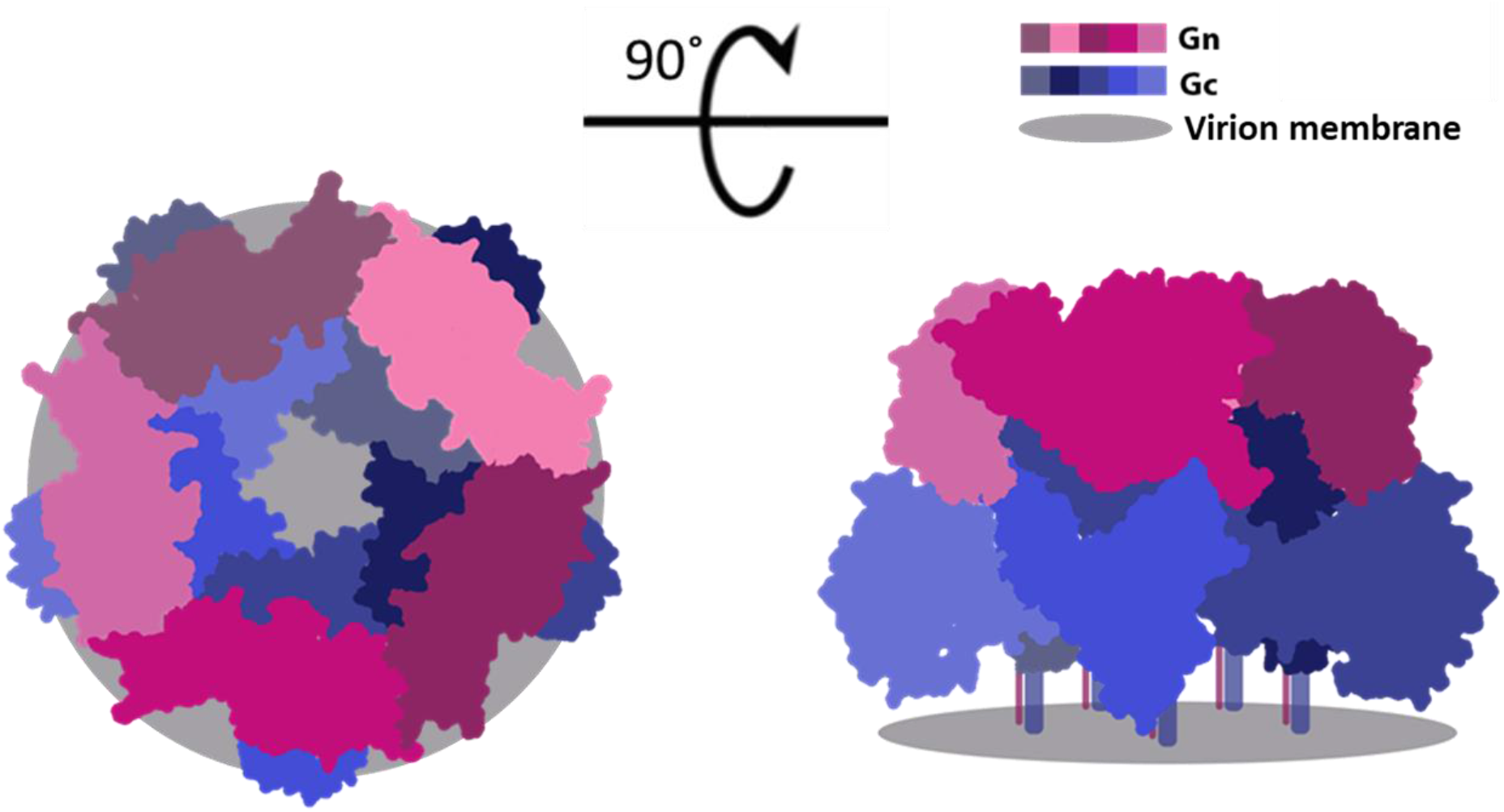
*Top (left) and side views (right) of a schematic surface representation of the pentameric assembly of envelope-displayed RVFV Gn-Gc heterodimers based upon integrative cryoEM and X-ray crystallography (PDB ID: 6F9F)(Halldorsson et al., 2018b) are shown. Pentamers and hexamers of Gn-Gc heterodimers encapsulate RVFV with T=12 icosahedral symmetry. Each membrane distally located RVFV Gn is rendered in a shade of purple and each membrane proximally located RVFV Gc is rendered in a shade of blue. The C-terminal regions of RVFV Gn and Gc are shown as lines due to the lack of high resolution structural information and are anchored to the virion membrane (grey)*.

While protection against RVFV infection has long been associated with the induction of nAbs targeting Gn and Gc (Dodd et al., 2013, Peters et al., 1988, LaBeaud, 2010), it can also arise from other mechanisms. Mice vaccinated with the immunodominant nucleoprotein (N), for example, show partial protection from RVFV challenge despite the absence of detectable nAbs (Lopez-Gil et al., 2013, Jansen van Vuren et al., 2011). Another study showed the cooperative effect of a non-neutralizing antibody improving protection in a mouse model (Gutjahr et al., 2020). Perhaps more surprisingly, mice vaccinated with modified Vaccinia Ankara (MVA) expressing both Gn and Gc failed to elicit nAbs but were still protected from viral challenge (Lopez-Gil et al., 2020). IgG1, the predominant subclass recognising the RVFV glycoproteins identified here, can efficiently trigger the classical route of complement (Vidarsson et al., 2014). While the contribution that non-neutralizing antibodies and cellular immunity have on protection against RVFV is less well characterized, numerous components of the immune response may function together during acute infection (including experimental challenge in animals) to provide protection. Studies on the immune response during acute RVFV infections in humans are needed to determine the role of other immune mechanisms in disease pathogenesis and protection.

Due to its extensive anti-viral properties, we have used IFNγ as a broad measure of the cellular response to RVFV in these individuals. The *ex vivo* ELISpot carried out here is a measure of effector memory T cells that are able to secrete IFNγ within hours of re-exposure to antigen (Calarota and Baldanti, 2013). We have shown that these cells, circulating in the peripheral blood, are relatively low in number in humans with historical RVFV infection. However, they are still detectable in the majority of individuals many years after exposure and they may potentially play an important role in protection. Recently, the T cell responses in human volunteers vaccinated with multiple doses of a formalin-inactivated RVF vaccine were characterized (Harmon et al., 2020). Despite also showing a generally low cellular response, IFNγ expression post-stimulation with Gn, Gc and N peptides were detected by ELISpot many years post vaccination. A previous study exploring the cellular response to RVFV in a non-human primate model have shown that non-lethal challenge was associated with early proliferation of CD4+ and CD8+ T cells in addition to early Th1 cytokine production, including IFNγ (Wonderlich et al., 2018). Another study in humans showed higher expression of IL-10, an anti-inflammatory cytokine, was associated with fatal cases (McElroy and Nichol, 2012). These findings, together with the critical role CD4+ T cells play in generating strong antibody responses (Dodd et al., 2013), indicate the importance and benefit of a robust cellular response.

Due to the presence of high-titer, long-lived nAbs, it is plausible that the naturally acquired immune response to RVFV may be sufficient for long-lasting protection, particularly as nAbs are associated with protection in animals. The observation that antibodies targeting Gn play a comparatively larger role in neutralization than those that target Gc has potential implications for vaccine design and is suggestive that the Gn may be the most critical antigen to include in RVFV candidate vaccines.

In summary, while our immunological assessments of individuals naturally exposed to RVFV provide useful data on immunity to infection, there is a lot that we do not know regarding their previous RVFV exposure; when or how often they became infected prior to sample collection, the route of exposure (mosquito bite vs contact with infected animal tissue) and their clinical manifestations. All of these factors will have likely influenced their subsequent response to RVFV and further work is needed to connect immunological attributes with clinical outcomes and disease severity. We have provided data characterizing the immune response in a large number of naturally exposed Kenyan adults who are likely protected from re-exposure. These data can provide a useful benchmark for immune responses elicited in future clinical trials of candidate vaccines.

## MATERIALS AND METHODS

### Study population

A cross-section of randomly selected households in Tana River County, Kenya (2013-2014) were enrolled after consenting and all human subjects within that household were sampled as part of a previous study estimating RVFV exposure (Bett et al., 2018). For this analysis, serum samples from 50 individuals were selected at random for antibody characterization from those identified as RVFV-exposed on the basis of seropositivity by both RVFV neutralization assay (VNT_50_ >10) and a diagnostic ELISA kit (BDSL, National Institute for Communicable Diseases, Centre for Emerging and Zoonotic Diseases, Johannesburg, South Africa) (Paweska et al., 2005). A further 7 RVFV-exposed individuals from the same study and with sufficient material available were assessed in the antibody depletion assays. Serum and peripheral blood mononuclear cells (PBMCs) were available from 200 adults under longitudinal surveillance for malaria studies in Kilifi County in 2018. For some of these adults, serum samples were available dating back to 1998 when they were enrolled into the longitudinal cohort. Additional screening of the serum samples collected in 2013-2014 identified positive individuals not sampled in 2018. Blood samples were collected in plain vacutainers and serum harvested after centrifugation at 3,000 RPM for 5 minutes, before storage at -80°C until use. Heparinised blood was used for PBMC isolation using standard protocols (Illingworth et al., 2013) and PBMCs cryopreserved until use. Ethical approval for this study was provided by the Kenya Medical Research Institute Scientific and Ethics Review Unit (SSC 3296), and the African Medical Research Foundation’s Ethics and Scientific Review Committee (approval number P65-2013).

### Protein Expression

#### RVFV Gn

cDNA of the Gn ectodomain (UniProt accession number P21401, residues 154-560) was cloned into pHLSec mammalian expression vector, which encodes a C-terminal hexa-histidine tag (Aricescu et al., 2006). Human embryonic kidney cells (HEK293T, ATCC CRL-1573) were transiently transfected and cell supernatants were purified by immobilized nickel-affinity chromatography (IMAC), using 5 mL HisTrap FF crude column and AKTA FPLC system (GE Healthcare). Gn protein was further purified by size exclusion chromatography (SEC) using a Superdex 200 10/300 Increase column (GE Healthcare) into 10mM Tris-HCl, 150mM NaCl, pH 8.0.

#### RVFV Gc

cDNA of the Gc ectodomain (UniProt accession number P21401, residues 691-1120) linked to an N-terminal SUMO tag, which also encodes a hexa-histidine tag, was subcloned into the pURD expression vector to generate a stable cell line in HEK293T cells (Seiradake et al., 2015). Cells were harvested after 5 days and RVFV Gc was purified from clarified cell supernatant by IMAC. The SUMO-tag was cleaved with 3C protease (Pearsons) overnight at room temperature. Cleaved RVFV Gc was purified from the SUMO-tag by IMAC.

The flowthrough containing Gc was further purified by SEC using a Superdex 200 10/300 Increase column (GE Healthcare) into 10mM Tris-HCl, 150mM NaCl, pH 8.0.

### Total IgG enzyme linked immunosorbent assays (ELISAs)

96 well NUNC flat bottom plates (ThermoFisher) were coated with 50 μl/well of Gn or Gc at a concentration of 1 μg/mL in carbonate bicarbonate buffer (Sigma) and incubated overnight at room temperature (RT). Plates were washed 6x with phosphate buffered saline containing 0.05% Tween20 (PBS/T) followed by blocking with 100 μl/well of casein (ThermoFisher) for 1 hour at RT. Plates were then tapped dry and test sera was diluted 1:100, 1:400 or 1:800 in casein. 50 μl/well was added to duplicate wells and incubated for 2h at RT. A positive reference serum made from a pool of high responders (using raw OD450 as measured by initial Gn/Gc ELISAs) was included on each plate as a standard curve. This was serially diluted in 2-fold steps from 1:100 to 1:51,200 in casein. Each dilution was added to the plate in duplicate, with the 1:100 dilutions given an arbitrary value of 10 RVFV antibody units (AU). Plates were washed as before and 50 μl/well of secondary antibody (goat anti-human whole IgG conjugated to HRP, Insight Biotechnology) was diluted 1:1,000 in casein and added to the plate for 1h at RT. Plates were washed a final time and developed by adding 100 μl/well of TMB developer (Abcam). After 8 minutes incubating in the dark at RT, 100 μl TMB stop solution (Abcam) was added. The optical density (OD) was read at 450 nm using a Varioskan flash reader. OD values were fitted to a 4-Parameter logistic model (Gen5 v3.09, BioTek) standard curve. An internal control of positive serum was included on every plate in duplicate made up from a 1:800 dilution of the positive standard. Test sera arbitrary units (AU) were calculated from their OD values using the parameters estimated from the standard curve.

### IgG avidity ELISAs

Total IgG avidity was determined by sodium thiocyanate (NaSCN) displacement, as described previously (Biswas et al., 2014). Briefly, sera was individually diluted in casein to normalise titers, according to their total IgG ELISA AU. Plate coating, blocking and development were performed the same as their respective total IgG ELISAs.

### IgG Subclass ELISAs

IgG subclass ELISAs were performed as per the total IgG ELISAs with the following exceptions: Secondary antibodies (biotin conjugated mouse anti-human IgG1, IgG2, IgG3 or IgG4 [Bio-Rad]) were incubated for 1h. After washing, 50 μL/well of eXtravidin-ALP (Sigma), diluted 1:5,000 in casein was added for 1h. Plates were washed and then developed by adding 50 μl/well of 4-nitrophenyl phosphate in diethanolamine buffer (Pierce). OD was read at 405 nm.

### Virus Neutralization assays (VNT and FRNT)

VNT: The dilution required to reduce neutralization by 50% (VNT50) was calculated, as described previously (Lopez-Gil et al., 2013). Briefly, sera pools were diluted in DMEM containing 10% FCS (D10) across a 96-well plate before addition of 100 TCID50 of RVFV MP-12 strain. Sera and virus were incubated at 37 °C for 1h in 200 μl before being added to a 96 well tissue culture (TC) plate containing a confluent monolayer of VERO cells. Plates were incubated at 37 °C for 72h before cells were fixed in PBS containing 10% formaldehyde for 1h and stained with PBS containing 1% crystal violet (Sigma) for 10 minutes. Plates were washed with water and left to dry before being scored by eye. Focus reduction neutralizing test (FRNT): This was carried out as previously described (Barsosio et al., 2019) with some alterations. Briefly, serum was diluted in 2-fold steps in triplicate from a starting dilution of 1:20.-Approximately 100 focus-forming units of RVFV (MP-12 strain) was added to each well and incubated for 1h at 37 °C before transferring mixture to 96-well Tissue Culture plate containing Vero E6 monolayers at 90% confluency. After 2h at 37 °C, virus/serum was removed and replaced with media. Plates were incubated a further 48h at 37 °C. Virus foci were detected by adding 1:1000 dilution of Anti-Gn 4D4 mouse mAb (bei resources, Cat. No. NR-43190) followed by 1:1000 dilution of horseradish peroxidase (HRP)-conjugated goat anti-mouse IgG antibody (Abcam, Cat. No. ab6789). After addition of 3,3’-diaminobenzidine (Sigma) substrate for 10 minutes at room temperature, the plates were washed a final time and air dried. Foci were counted using an AID ELISpot reader. Serum dilution required to reduce virus foci by 50% compared to virus-only control wells (6 replicates) were calculated from a 4-parameter standard curve using GraphPad Prism version 8 (GraphPad Software Inc., California, USA).

### Gn and Gc specific antibody depletion

In order to deplete anti-Gc and/or anti-Gn IgG, 96-well NUNC ELISA plates were coated with 5 μg/mL of recombinant Gn or Gc or carbonate-bicarbonate buffer alone. Plates were incubated overnight. After washing with PBS/T as previously described, plates were blocked with 200 μL/well of 1% casein in PBS (Sigma) for 2h. Neat pooled human sera was diluted 1:50 in DMEM containing 2% L-glutamine and 1% penicillin/streptomycin (D0) with subsequent 1:2 titrations up to 1:102400. Diluted sera was added to each antigen-coated plate (including the blocked but uncoated control plate) and incubated on a plate rocker at 10 RPM for 4h at room temperature. Serum was then removed and the depletion step repeated once more on additional coated and blocked plates. For the dual Gn/Gc depletion, serum was depleted twice on Gn-coated plates and twice on Gc-coated plates. Reduction of antigen-specific antibodies after final depletion was confirmed by ELISA. Serum was diluted 1:2 with Casein and total IgG ELISA was carried out as described. The endpoint titer is defined as the interpolated dilution (OD at 450nm versus dilution of serum) at which the OD of the sample is 0.15. The remaining serum was added to the VNT plate to determine nAb titer, as previously described, with final serum concentrations ranging from 1:200-1:102,400 (Barsosio et al., 2019).

### *Ex-vivo* IFNγ ELISpot

264 15mer peptides overlapping by 11 amino acids, covering the Gn-Gc polyprotein (Genbank accession number DQ380208, residues 132-1197) were synthesised (Mimotopes). Peptides were split into 5 pools of 52/53 peptides each at 1.25 μg/mL final concentration. ELISpots were carried out as previously described (Kimani et al., 2014). Responses from unstimulated wells were subtracted before quantifying the response to the 5 pools and summing them. Positive control wells were stimulated with SEB at 0.02 μg/mL.

### Statistical analysis

Statistical analysis was carried out using GraphPad Prism version 8 (GraphPad Software Inc., California, USA). All analyses of correlations were performed using non-parametric Spearman’s correlation tests using a two-sided p value < 0.05 as the cut-off for statistical significance. Comparisons between two groups were analysed using Kruskal-Wallis test or unpaired T-test.

## Data Availability

The data in this manuscript is not available elsewhere

## AUTHOR CONTRIBUTIONS

Conceptualization D.W., G.M.W. and T.A.B.; Methodology, D.W., E.R.A., J.N.G., H.K.K. G.M.W; Investigation, D.W., M.H.A.C., J.N.G., H.K.K.; Resources, R.H., I.T., S.B., E.R.A., D.M., J.M., B.B.; Writing - Original Draft, D.W; Writing - review editing, G.M.W., T.A.B.; Funding acquisition, G.M.W., T.A.B; Supervision; G.M.W., T.A.B.

## ACKNOWLEDGEMENTS

This work was supported through grants from the Wellcome Trust (grants no. 203077/Z/16/Z and 203141/Z/16/Z), an Oak foundation fellowship to GMW, and by the Medical Research Council (MR/L009528/1m MR/S007555/1, MR/N002091/1 to T.A.B.). This manuscript was submitted for publication with permission from the Director of the Kenya Medical Research Institute.

